# Character-Level Linguistic Biomarkers for Precision Assessment of Cognitive Decline: A Symbolic Recurrence Approach

**DOI:** 10.1101/2025.06.12.25329529

**Authors:** Kevin Mekulu, Faisal Aqlan, Hui Yang

## Abstract

Early and accurate detection of Alzheimer’s disease (AD) remains a critical challenge for precision health. Traditional cognitive assessments often miss subtle, individualized patterns of decline, while conventional linguistic analyses focus on word-level features that may overlook fine-grained speech disruptions. We test the hypothesis that character-level features in speech transcripts capturing pauses, repetitions, and hesitations at the finest linguistic granularity can serve as novel biomarkers for cognitive decline, revealing personalized linguistic signatures that manifest uniquely in each individual. Our biomarker discovery framework employs symbolic character-level encoding followed by recurrence quantification analysis to transform speech transcripts into visual recurrence plots that reveal temporal speech dynamics. Siamese networks learn embeddings from these plots to capture discriminative patterns at the character level. We validate our hypothesis using the DementiaBank corpus, demonstrating that character-level biomarkers achieve superior discriminative capability compared to conventional word-level approaches (95.9% vs. 87.5% AUC), while providing interpretable recurrence plot visualizations. Our findings establish that character-level linguistic features contain significant biomarker information for cognitive assessment, representing a fundamental shift from word-based to character-based analysis for precision health applications in dementia screening.

## I. Introduction

**A**Lzheimer’s disease (AD) and related neurocognitive disorders represent a growing global health crisis, affecting millions of individuals and placing immense burdens on healthcare systems [1]. Early and accurate detection of cognitive decline is essential for precision health, enabling targeted interventions, timely support, and individualized care strategies that improve quality of life and long-term outcomes [2], [3].

Traditional cognitive assessments, such as the Mini-Mental State Examination (MMSE) and Montreal Cognitive Assessment (MoCA), remain the clinical standard but are limited by subjectivity, interviewer and educational bias, and a tendency to identify dementia only at later stages [4]–[8]. As a result, many cases of cognitive impairment go undetected or are diagnosed too late for optimal intervention.

Recent advances in artificial intelligence (AI) and natural language processing (NLP) have enabled the detection of subtle linguistic and speech patterns associated with dementia [9], [10]. However, many AI-based approaches operate as black boxes, providing limited transparency or interpretability for clinicians and patients, and often failing to deliver individualized, actionable insights required for precision health applications [11].

A key challenge for next-generation dementia screening is not only robust early detection but also interpretability and the ability to capture fine-grained, individual differences. Character-level symbolic analysis, in contrast to standard word- or token-level models, can detect subtle speech disruptions, repetitions, and disfluencies that may serve as early and personalized indicators of cognitive decline [12], [13]. Moreover, interpretable representations of these linguistic features can facilitate more tailored risk assessment and clinical decision support [14], [15].

In this work, we test the hypothesis that character-level features in speech transcripts can serve as effective biomarkers for cognitive decline. We present a methodological framework that integrates recurrence quantification analysis (RQA) of character-encoded speech with deep metric learning to capture the subtle linguistic disruptions that may precede more obvious semantic or syntactic deficits. By transforming character sequences into recurrence plots and using Siamese networks to learn similarity metrics, our approach aims to identify patient-specific signatures of cognitive change at this fine-grained linguistic level. Figure 1 illustrates how speech transcripts can be transformed into recurrence plots that reveal these character-level patterns.

**Fig. 1:**
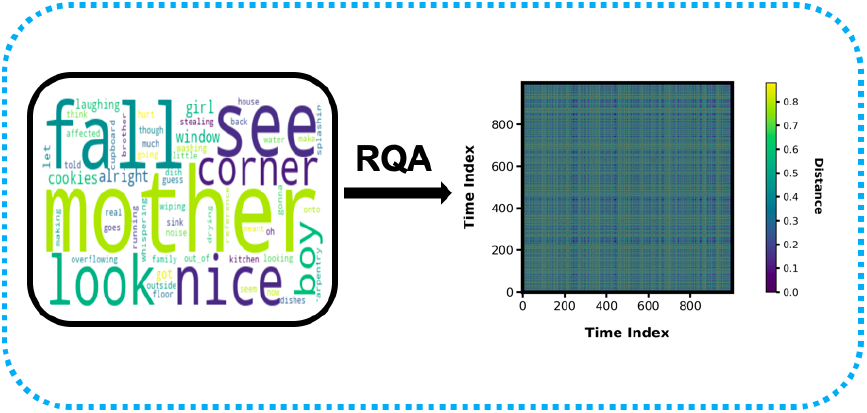
Illustration of a recurrence plot generated from a speech transcript.

We validate our character-level biomarker approach by benchmarking against traditional word-level models, demonstrating the discriminative power of these fine-grained linguistic features. We discuss both the potential and the practical limitations of character-level biomarkers within the broader landscape of precision health for dementia care, and explore how these individualized linguistic signatures could support personalized screening and monitoring.

### Paper Structure

Section II reviews related work and the precision health context. Section III details our methodology. Section IV presents experimental results, benchmarking and discusses implications for individualized care. Section V concludes with future directions.

## II. Research Background

Dementia, a progressive disorder impacting neurocognitive function, disrupts memory, reasoning, and communication, ultimately leading to severe disability and mortality. Alzheimer’s disease (AD) accounts for 60–80% of dementia cases [16]. Early-stage identification is critical for optimizing prognosis and deploying individualized interventions, yet remains an unmet clinical need due to the limitations of conventional diagnostic tools.

Traditional assessments—clinical interviews and paper-based tests—are often subjective, time-consuming, and susceptible to cultural and educational biases. Critically, they frequently miss the subtle, early-stage linguistic or behavioral changes that precede overt cognitive decline [17].

Recent advances in artificial intelligence (AI) and deep learning (DL) have opened new avenues for noninvasive, data-driven diagnostics in neurocognitive disorders. Most research has centered on leveraging DL for neuroimaging [18] or retinal data [19], but there is a rapidly growing focus on speech and language as rich biomarkers for dementia and related conditions.

Nonlinear speech dynamics, such as disruptions in fluency, repetition, or lexical diversity, are recognized as sensitive early indicators of cognitive impairment [20]. Orozco-Arroyave et al. [21] demonstrated that nonlinear analyses can capture disease-relevant speech patterns, achieving promising accuracy in neurological disorders. Symbolic recurrence, a method rooted in nonlinear dynamics, was previously applied to speech data in our earlier preprint to identify temporal structure in linguistic patterns [22]. In contrast to our earlier work, the present study directly benchmarks symbolic recurrence against traditional TF-IDF features popularized by Fraser et al. [23], highlighting significant gains in sensitivity.

Convolutional neural networks (CNNs) and deep architectures, such as VGG19 [24] and autoencoders [25], have achieved high accuracy in classifying AD from various input modalities. However, these models often act as black boxes, providing limited interpretability or individualized insights needed for clinical adoption in precision health [26]. Recent work highlights the importance of transparent, explainable AI methods that can provide actionable, person-specific information to support clinicians and patients [27], [28].

Furthermore, in our previous study, we introduced a novel character-level Markov modeling approach, termed *CharMark*, which demonstrated that steady-state character transitions could effectively distinguish between healthy and impaired individuals [29]. We also conducted a comparative study of large language models (LLMs) and pre-trained encoders like BERT for dementia screening, finding that simpler encoder-based models often outperform LLMs on structured tasks like Cookie Theft picture descriptions [30].

Our approach builds on these advances by integrating deep metric learning (Siamese networks [31]–[33]) with recurrence quantification analysis (RQA) of character-encoded speech transcripts [9], [10]. RQA captures the nonlinear temporal structure of speech, while deep metric learning learns discriminative, interpretable representations of individual linguistic patterns. This enables both robust biomarker identification and nuanced, personalized insights—supporting the broader precision health agenda.

By analyzing character-level features through symbolic encoding, RQA visualization, and deep metric learning, our methodology tests the hypothesis that subtle linguistic markers at the character level can serve as interpretable, personalized biomarkers for cognitive decline—potentially offering earlier and more individualized detection than traditional word-level analyses or black-box AI approaches.

## III. Research Methodology

Our methodology tests the hypothesis that individual character-level features in speech transcripts can serve as biomarkers for early dementia detection. We integrate symbolic character-level encoding, recurrence quantification analysis (RQA), deep metric learning with Siamese networks, and tree-based classification as analytical tools to evaluate this hypothesis. The overall pipeline is shown in Fig. 2.

**Fig. 2:**
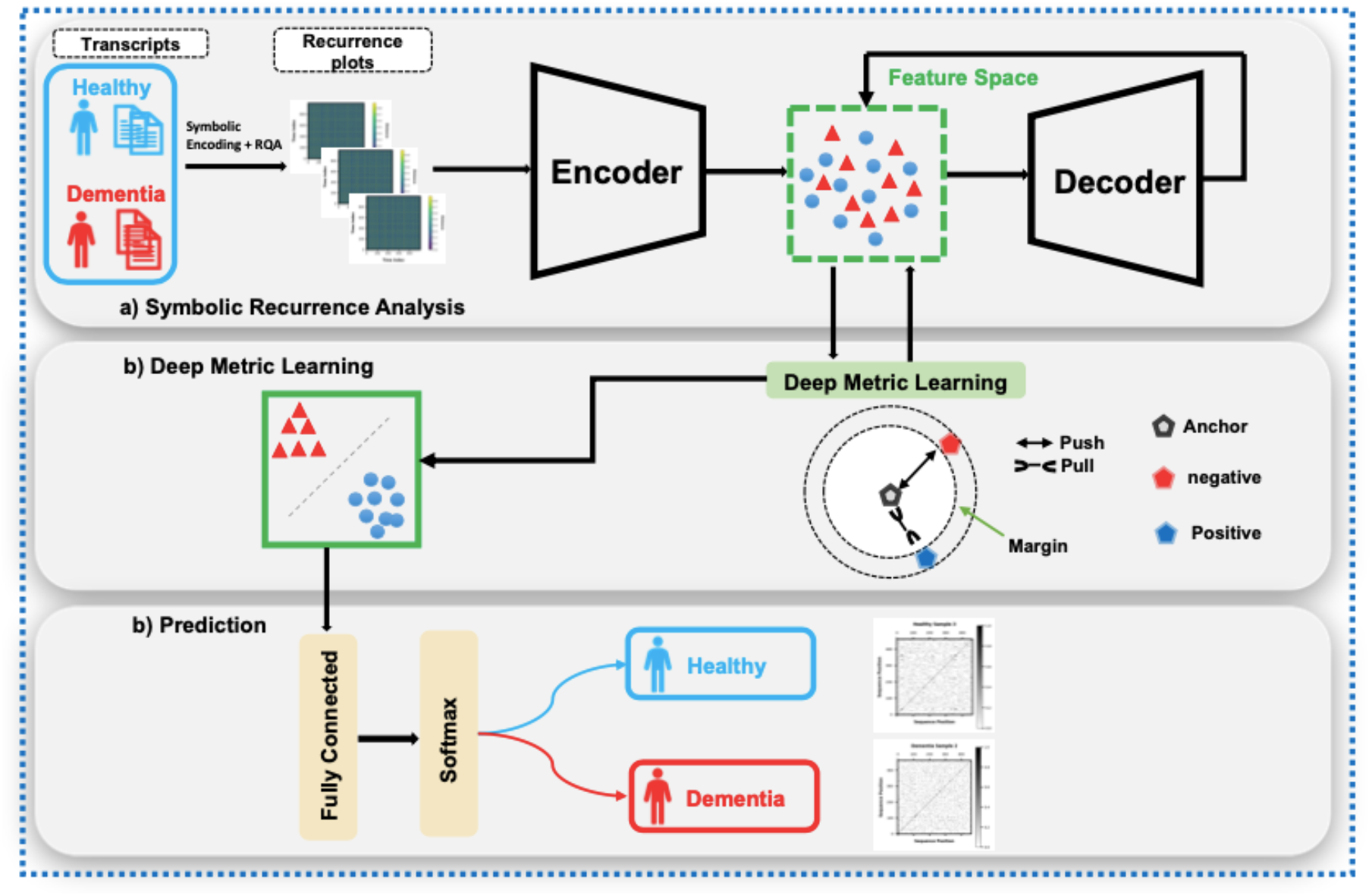
Overview of the proposed methodology for speech-based dementia detection.

### A. Design Rationale and Hypothesis Formulation

Our primary hypothesis is that character-level features from speech transcripts can serve as effective biomarkers for cognitive decline, potentially offering finer-grained indicators than traditional word or sentence-level analyses. This hypothesis is investigated through a methodological pipeline that preserves and analyzes the character-level structure of speech.

#### Why Character-Level Encoding as Biomarkers?

Character-level features (e.g., pauses, repetitions, hesitations) reflect fine-grained language changes that precede more severe semantic or syntactic deficits [34], [35]. These microlinguistic elements manifest early in cognitive decline and may serve as sensitive biomarkers that clinicians already intuitively assess during cognitive examinations.

#### Why Text Transcripts Instead of Audio?

While acoustic features (e.g., prosody, vocal quality, speech rate) provide valuable information about cognitive status [36], [37], text-based analysis offers several distinct advantages for biomarker development. Text transcripts provide a robust, scalable, and de-identifiable modality that can be retrospectively applied to existing clinical records. Linguistic features from transcripts have been shown to be sensitive indicators of cognitive decline [23], [38] and are more easily standardized across clinical settings, reducing variability from recording conditions, microphone quality, and background noise that often affect acoustic analyses.

Furthermore, transcript-based approaches facilitate easier integration with existing clinical workflows where recording quality may be inconsistent. Our character-level encoding preserves many paralinguistic features typically lost in word-level analyses (e.g., pauses, hesitations, etc) while maintaining the practical advantages of text-based assessment. This approach also allows us to focus specifically on linguistic disruptions rather than potential confounds from non-cognitive factors that affect voice quality, such as respiratory conditions or dentition issues common in older adults [39].

#### Why Recurrence Quantification Analysis (RQA)?

RQA captures nonlinear, temporal recurrences in linguistic structure at the character level—revealing disruptions in speech fluency, repetition, and fragmentation typical of early dementia, beyond the reach of traditional NLP metrics [9], [10].

#### Why Machine Learning for Biomarker Validation?

Our machine learning pipeline serves primarily to validate whether character-level features indeed contain sufficient discriminative information to function as biomarkers. Siamese networks and XGBoost are used not just for classification but to confirm whether the character-level encoding hypothesis has merit, with performance metrics serving as validation measures of our biomarker approach.

### B. Experimental Data

We employ the DementiaBank Pitt Corpus, comprising 552 transcripts collected using the Cookie Theft picture description task. Transcripts were transcribed according to CHAT standards [40]. Our analysis includes 168 Alzheimer’s disease (AD) patients (194 transcripts) and 98 healthy controls (242 transcripts). Table I summarizes participant demographics.

**TABLE I:** Demographic Information of Participants (DementiaBank Pitt Corpus)

|  | Healthy Control (HC) | Alzheimer's Disease (AD) |
| --- | --- | --- |
| No. of Participants | 98 | 168 |
| Gender (M/F) | 31/67 | 55/113 |
| Age (mean ± SD) | 64.7 ± 7.6 | 71.2 ± 8.4 |
| Education (years) | 14.0 ± 2.3 | 12.2 ± 2.6 |
| MMSE Score | 29.1 ± 1.1 | 19.9 ± 4.2 |

### C. Character-Level Encoding as Potential Biomarkers

To test our hypothesis that character-level features can serve as biomarkers, we first removed all interviewer utterances from the transcripts to focus exclusively on patient speech patterns. We then implemented a character-level symbolic encoding approach that preserves the fine-grained temporal structure of speech production.

Each character in the transcript—including alphabetic characters, punctuation marks, and whitespace—was mapped to a unique integer code (e.g., ‘a’ → 1, ‘b’ → 2, …, ‘ ‘ → 27, ‘.’ → 28, etc.). This encoding preserves potential biomarkers of cognitive function: spaces between words capture hesitation and pausing patterns, punctuation marks reflect prosodic boundaries and speech organization, and character sequences reveal potential disruptions in articulation and word-finding difficulties. By treating each character as a potential biomarker, we capture subtle linguistic disruptions that may occur before more obvious semantic and syntactic deficits emerge.

All encoded sequences were subsequently standardized to uniform length through padding or truncation to ensure consistent processing across samples. Specifically, we determined the maximum sequence length across all transcripts in the dataset and applied post-padding (adding zeros to the end of shorter sequences) and post-truncation (removing characters from the end of longer sequences) to achieve uniform dimensionality. This padding strategy preserves the temporal ordering of character-level features while ensuring that the beginning of each transcript which typically contains the most informative speech patterns remains intact. Zero-padding at sequence ends maintains the natural flow of speech production without introducing artificial patterns that could confound the analysis.

This standardization approach maintains the temporal dynamics of speech production while focusing specifically on character-level features as potential biomarkers for cognitive decline. The uniform sequence length enables batch processing through the Siamese network architecture while preserving the character-level temporal structure that captures subtle linguistic disruptions such as hesitations, repetitions, and pausing patterns.

### D. Recurrence Quantification Analysis (RQA)

To visualize the temporal dynamics of character-level features, encoded sequences are transformed into recurrence plots using RQA. A recurrence plot is defined by the matrix:

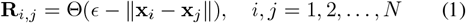

where Θ is the Heaviside step function, *ϵ* is the threshold parameter (set dynamically as 0.1*×* sequence standard deviation), and **x**_*i*_ and **x**_*j*_ are the encoded states at positions *i* and *j* in the character sequence.

This transformation converts the linear character sequences into two-dimensional recurrence plots that reveal the underlying patterns of repetition, determinism, and complexity in the speech samples. These recurrence plots serve as rich visual representations of speech dynamics, potentially capturing subtle differences between healthy and impaired cognitive states that might not be evident in the raw character sequences.

Unlike traditional approaches that extract specific RQA metrics (such as determinism or recurrence rate), we utilize the entire recurrence plot as a holistic representation of speech dynamics. This preserves all the structural information in the nonlinear patterns for subsequent deep learning analysis.

Example recurrence plots for dementia and control cases are shown in Fig. 3. Visual inspection reveals distinct patterns between the groups, with dementia subjects typically showing more fragmented recurrence structures.

**Fig. 3:**
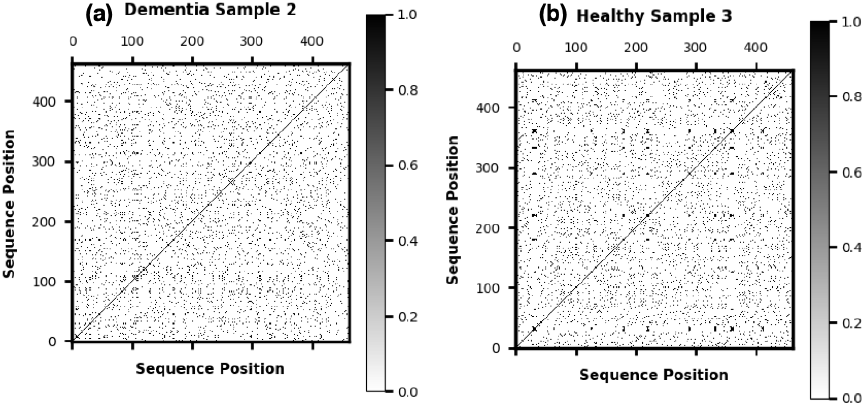
Sample recurrence plots: (a) dementia subject, (b) healthy subject.

### E. Deep Metric Learning with Siamese Networks

To validate whether our character-level features can effectively distinguish between cognitive states, pairs of recurrence plots are directly input to a Siamese convolutional neural network without extracting intermediate features. The Siamese architecture consists of two identical convolutional neural networks with shared weights that process pairs of inputs, projecting them into an embedding space where the distance between embeddings reflects similarity.

Prior to network input, the recurrence plots undergo preprocessing to ensure consistent dimensionality and scale. Each recurrence plot is resized to a fixed dimension of 128 *×* 128 pixels, preserving the structural patterns while standardizing the input size. The pixel values are then normalized to the range [0, 1] to improve training stability and convergence. This preprocessing step ensures that the network focuses on the structural patterns in the recurrence plots rather than absolute intensity values.

The Siamese network architecture consists of identical twin subnetworks that share weights, as detailed below:

#### Algorithm 1 Siamese Network Architecture

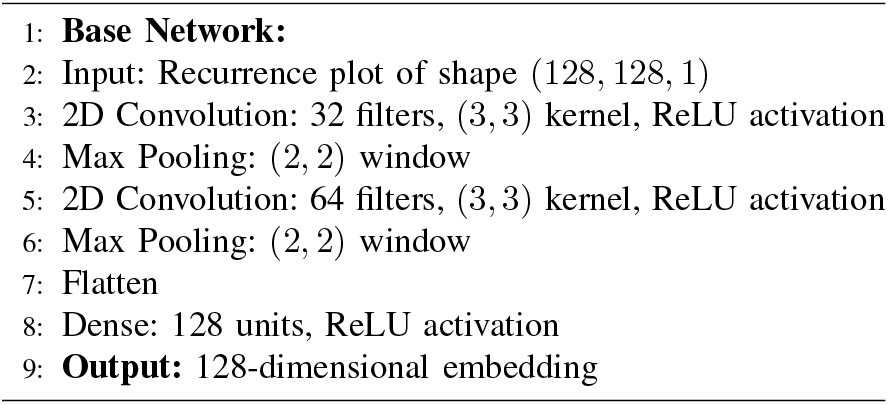

The network is trained using contrastive loss:

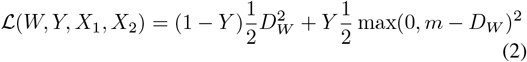

where *Y* is the binary label (1 for different class pairs, 0 for same class), *D*_*W*_ is the Euclidean distance between the embeddings in the learned space, *m* is the margin parameter (set to 1.0) that defines the minimum separation between classes, and *W* represents the network parameters. This loss function encourages similar samples to cluster together while pushing dissimilar samples apart by at least the margin *m*.

The model is optimized using Adam optimizer with a learning rate of 0.001. During training, we randomly sample pairs of recurrence plots with equal proportions of same-class and different-class pairs to ensure balanced learning. The resulting 128-dimensional embeddings capture the discriminative characteristics of speech patterns at the character level.

The Siamese network learns to extract discriminative features directly from the recurrence plots that can effectively differentiate between healthy and dementia-affected speech patterns. By letting the deep network learn relevant features automatically from the raw recurrence plots, we avoid potential information loss that might occur when reducing the plots to predefined metrics. The quality of these learned embeddings serves as a measure of how effectively character-level features can function as biomarkers.

### F. Biomarker Validation through Machine Learning

To validate our hypothesis that character-level features can serve as biomarkers, we input the learned embeddings to an XGBoost classifier. The performance of this classifier provides evidence for whether character-level features contain sufficient discriminative information to function as reliable biomarkers of cognitive decline. The XGBoost algorithm constructs an ensemble of decision trees through gradient boosting, with the objective function:

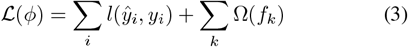

where *l* is the loss function measuring the difference between the prediction *ŷ*_*i*_ and the true label *y*_*i*_, and Ω is a regularization term that penalizes the complexity of the model, preventing overfitting.

We employed an extensive grid search to optimize the XGBoost classifier hyperparameters, including learning rate, maximum tree depth, minimum child weight, subsample ratio, and regularization parameters (alpha and lambda). The optimal hyperparameter configuration was selected based on cross-validation AUC score.

### G. Evaluation and Benchmarking

To evaluate the effectiveness of our character-level biomarker approach, we benchmark it against a widely-used traditional word-level method. The baseline model employs Term Frequency-Inverse Document Frequency (TF-IDF) vectorization followed by a logistic regression model.

The TF-IDF approach represents each transcript as a high-dimensional vector where:

- Each dimension corresponds to a unique word in the vocabulary
- The value for each word is proportional to its frequency in the document
- Words that appear in many documents are downweighted to reduce the importance of common words

This word-level baseline focuses on lexical content (the specific words used) rather than the fine-grained paralinguistic features captured by our character-level approach. It represents the conventional text classification methodology that has been widely used in previous dementia detection studies [23], [41].

By comparing our character-level biomarker approach against this word-level baseline, we aim to determine whether the subtle linguistic patterns captured by character-level encoding provide additional discriminative information beyond traditional lexical features. Both models are evaluated using Area Under the ROC Curve (AUC), with results presented in Section IV. This comparison directly tests our hypothesis that character-level features can serve as effective biomarkers for cognitive decline.

### H. Experimental Design

Data is split 80/20 (train/validation) with stratified sampling. Five-fold stratified cross-validation is used to ensure robust performance estimation. All parameter settings, data splits, and code details are reported for reproducibility and will be released upon publication, in accordance with journal policies.

## IV. Experimental Results and Validation

Our experimental results focus on validating whether character-level features in speech transcripts can serve as effective biomarkers for cognitive decline. We evaluate this hypothesis through a series of experiments comparing our character-level approach against traditional word-level models, assessing both discriminative power and consistency.

### A. Character-Level Approach Performance

We evaluated our character-level biomarker approach using stratified 5-fold cross-validation to ensure robust assessment across different data subsets. Figure 5 presents the ROC AUC scores for each fold, revealing strong and consistent performance with a mean AUC of 95.9%. This high discriminative ability suggests that character-level features indeed contain significant information relevant to cognitive status.

**Fig. 4:**
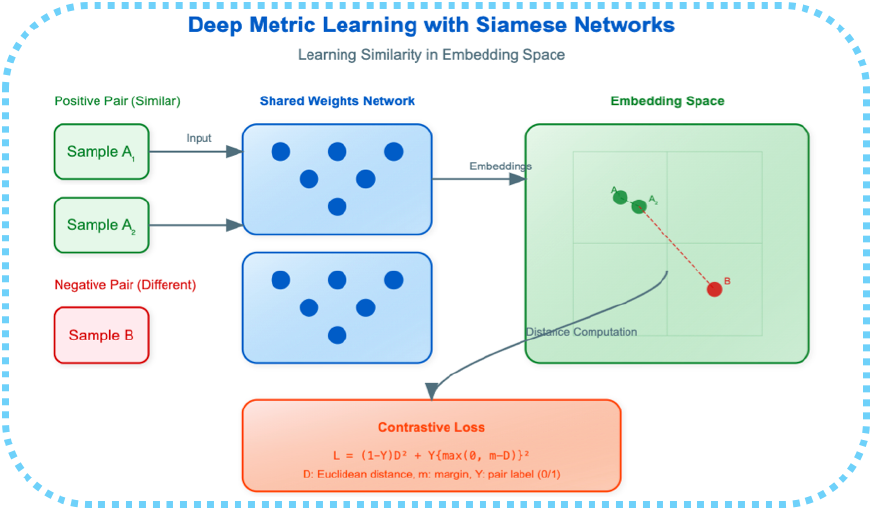
Siamese network architecture for learning patient-specific speech embeddings.

**Fig. 5:**
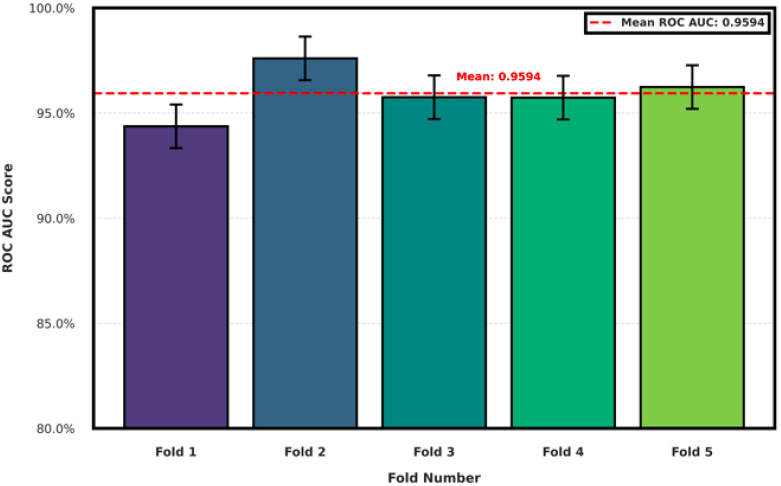
ROC AUC scores across stratified 5-fold cross-validation for our character-level biomarker approach. The mean AUC of 95.9% demonstrates strong and consistent performance.

### B. Comparison with Word-Level Approaches

To contextualize our findings, we compared our character-level approach against a traditional word-level model using TF-IDF vectorization with logistic regression—a widely used baseline in clinical text classification [23], [41]. This comparison directly tests whether character-level features offer advantages over conventional word-level analysis.

As shown in Figure 6, the word-level approach achieved a mean AUC of 87.5% across the same 5-fold stratified cross-validation. While this performance is reasonable, our character-level approach showed an absolute improvement of 8.4 percentage points, representing a substantial gain in discriminative ability.

**Fig. 6:**
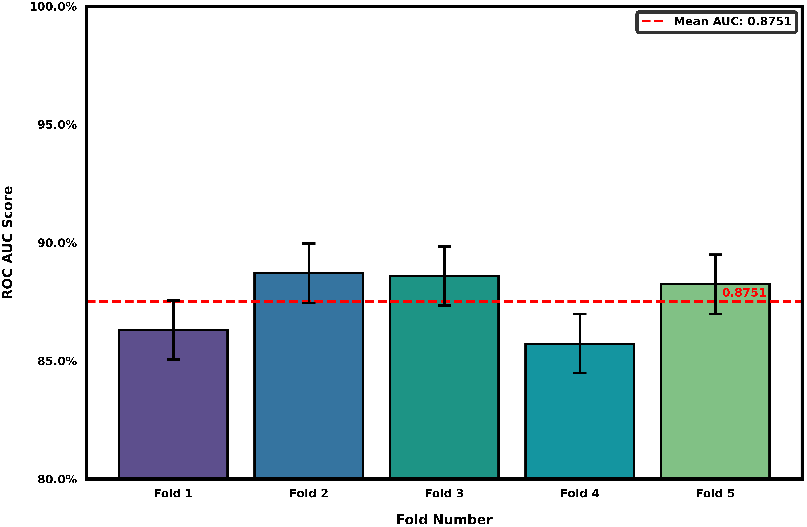
ROC AUC scores for the baseline TF-IDF + Logistic Regression model across 5-fold cross-validation. The mean AUC of 87.5% is notably lower than our character-level approach.

Table II provides a detailed comparison of both approaches. Beyond the higher mean performance, our character-level approach also demonstrated excellent consistency across folds, with similar standard deviation to the word-level model but a higher minimum AUC. This suggests that character-level features provide more robust biomarkers across different subsets of subjects.

**TABLE II:** Performance Comparison of Character-Level vs. Word-Level Approaches.

| Model | Mean AUC | Std Dev | Min AUC | Max AUC |
| --- | --- | --- | --- | --- |
| Character-level (Ours) | 95.9% | 1.2% | 94.3% | 97.8% |
| Word-level (TF-IDF) | 87.5% | 1.2% | 85.7% | 88.9% |

### C. Interpretability and Clinical Relevance

While quantitative performance is important, the primary contribution of this research is demonstrating that character-level features can serve as interpretable biomarkers for cognitive decline. The recurrence plots generated from character-level encoding provide visual representations of speech dynamics that clinicians can potentially inspect, offering an interpretable window into the subtle disruptions in language production that may occur before more obvious linguistic deficits emerge.

The superior performance of our character-level approach over word-level analysis supports our hypothesis that fine-grained linguistic patterns at the character level contain valuable biomarker information that may be missed by conventional word-level approaches. These patterns—including hesitations, repetitions, and disfluencies represented by spaces and punctuation—appear to capture fundamental aspects of cognitive processing that manifest in speech production.

This finding has significant implications for precision health approaches to dementia care, suggesting that individualized assessment of character-level linguistic patterns could potentially enable earlier detection and more personalized monitoring of cognitive changes than approaches focused solely on lexical content.

## V. Conclusions

In this study, we tested the hypothesis that character-level features in speech transcripts can serve as effective biomarkers for cognitive decline. Our results provide strong support for this hypothesis, demonstrating that fine-grained linguistic patterns captured through character-level encoding, recurrence quantification analysis, and deep metric learning offer substantial discriminative power for assessing dementia-related changes in speech.

The performance of our character-level approach (mean AUC 95.9%) significantly outperformed the traditional word-level TF-IDF model (mean AUC 87.5%), highlighting the value of analyzing these more subtle linguistic features. This performance gap suggests that important indicators of cognitive status may be embedded in the paralinguistic aspects of speech such as pauses, repetitions, and hesitations that are preserved through character-level encoding but missed by conventional word-level analyses.

Beyond classification performance, our approach offers several advantages for precision health applications. The recurrence plots generated from character-level sequences provide interpretable visualizations that could potentially help clinicians understand individual speech patterns. The patient-specific embeddings learned by the Siamese network may capture personalized linguistic signatures of cognitive status, supporting individualized assessment and monitoring.

While promising, our approach has certain limitations. The current study was conducted on a single dataset with controlled picture description tasks, and future work should validate these findings across more diverse linguistic contexts and spontaneous speech. Additionally, longitudinal studies would be valuable to assess how these character-level biomarkers evolve during disease progression and whether they can detect changes before conventional clinical assessments.

Future directions include exploring the relationship between specific character-level patterns and particular cognitive domains, developing more accessible implementations for clinical use, and investigating whether similar approaches could be applied to other neurological and psychiatric conditions with linguistic manifestations.

In conclusion, our findings establish character-level features as promising biomarkers for cognitive decline, with potential applications in early screening, monitoring, and personalized intervention planning. By capturing subtle disruptions in speech production at this fine-grained level, our approach contributes to the growing toolkit of precision health methods for dementia care offering a noninvasive, interpretable window into cognitive function that complements existing clinical assessments.

## Data Availability

The data that support the findings of this study are available from the Pitt Corpus within DementiaBank, maintained by TalkBank at Carnegie Mellon University and the University of Pittsburgh School of Medicine. Access to the data in DementiaBank is password protected and restricted to members of the DementiaBank consortium group. In accordance with TalkBank rules, any use of data from this corpus must be accompanied by appropriate corpus references and acknowledgment of grant support (NIA AG03705 and AG05133). Interested researchers can request access to the Pitt Corpus by visiting https://dementia.talkbank.org/access/English/Pitt.html. Established researchers and clinicians working with dementia can contact TalkBank to request access credentials.

## Acknowledgment

The authors of this work would like to acknowledge the NSF grants IIS-2302834 and MCB-1856132 for funding this research.Any opinions, findings, or conclusions found in this paper originate from the authors and do not necessarily reflect the views of the sponsor.

